# Improvement in Sleep Duration was Associated with Higher Cognitive Function among Middle-Aged and Elderly Chinese Participants

**DOI:** 10.1101/2020.04.09.20060277

**Authors:** Jianian Hua, Hongpeng Sun, Yueping She

## Abstract

**Study objectives:** Rodent studies suggested that improvement in sleep duration might correlate with better cognitive function. We aimed to examine the associations between changes in sleep duration and cognitive function.

**Methods:** 10325 individuals from the China Health and Retirement Longitudinal Study (CHARLS) were included. Self-reported nocturnal sleep duration and cognitive function were assessed in CHARLS 2011, 2013 and 2015 (Wave 1, Wave 2, Wave3). Cognitive function was assessed by a global cognition score, which included three domains: episodic memory, figure drawing and Telephone Interview of Cognitive Status (TICS). Generalized additive models (GAM) and Generalized estimation equations (GEE) were used to examine the associations between baseline sleep duration and longitudinal cognitive function. We used generalized linear models (GLM) to study the associations between changes in sleep duration and cognitive function in Wave 3.

**Results:** After adjusting for potential confounders, change from short sleep duration (SSD) to moderate sleep duration (MSD) was associated with better global cognition scores (β=0.54, P <0.01). Change from SSD to long sleep duration (LSD) (β=-0.94, P <0.001) or change from LSD to SSD (β=-1.38, P <0.01) was associated with lower global cognition. For individuals with MSD, ≥2 h increase (β=-0.89, P <0.001) or decrease (β=-0.70, P <0.001) in sleep duration was associated with lower global cognition.

**Conclusions:** For short sleepers, improvement in sleep duration correlated with better cognition. For long sleepers, there was no need to reduce sleep duration. Excessive changes or deviation from the moderate duration was associated with lower cognition.

## 1 Introduction

The World Health Organization estimated that about 40 million people were living with dementia in 2015, and the number would increase 10 million per year^1^. Cognitive function test is a key way to detect dementia. Since dementia causes irreversible effect on life quality and strain on global health, it is imperative to find modifiable risk factors for lower cognitive function so that we can prevent or postpone dementia.

Considerable studies have found associations between sleep and cognitive function^2^. A recent meta-analysis revealed an inverted-U shaped association between sleep duration and risk of Alzheimer’s disease^3^. In the result, compared with people with moderate sleep duration, both people with short sleep duration and long sleep duration had higher risk.

Experimental studies have showed associations between sleep, circadian rhythm, and neurodegenerative diseases, especially for Alzheimer’s disease^4,5^. Under the guidance of this association, researchers pointed out that improvement in sleep duration might be a therapeutic target in dementia treatment or prevention^5-7^. Few studies have examined the effect of change in sleep duration on cognition. Limited by sample sizes or methodology, all their conclusions were that increased or decreased sleep duration was associated with lower cognitive function or higher risk of dementia^8-10^. To data, no epidemiology or clinical study proved that improvement in sleep duration correlated with better cognitive function. Moreover, the association between changes in sleep duration and cognition have yet to be examined among middle-aged Chinese.

Considering the “inverted-U shaped association” mentioned above, we hypothesized that a change towards the optimal sleep duration was associated with better cognitive function while deviation from the optimal was linked to lower cognitive function. Here, a large-scale population-based prospective study, the China Health and Retirement Longitudinal Study (CHARLS), would enable us to verify our hypothesis. The aim of our study was to: (1) re-examine the inverted-U shaped association between sleep duration and cognitive function among Chinese participants; (2) investigate the longitudinal association between baseline sleep duration and cognitive function over a period of time; (3) study the association between change in sleep duration and cognitive function.

## 2 Methods

### 2.1 Study Sample

Participants included in our study were from the China Health and Retirement Longitudinal Study (CHARLS), which was a prospective survey designed to provide scientific research related to the aged in China. The participants were non-institutionalized, aged 45 years and older and from 28 provinces in China. Baseline survey were conducted on 17708 participants between 2011 and 2012. The survey was conducted and followed up every two years^11^.

In the CHARLS, a total number of 15700 participants had data on sleep duration and cognitive tests in 2011 (Wave 1). 104 individuals under 45 years old till 2015 were excluded. 380 individuals with a history of brain damage or mental retardation were excluded. Among the rest 15216 participants, 12807 (84.2%) of them were followed up and had complete data on sleep duration and cognitive tests in 2013 (Wave 2). 10325 (80.6%) participants were followed up and had relative data in 2015 (Wave 3). The selection diagram and criteria for exclusion are provided in **Figure 1**.

**Figure 1.**
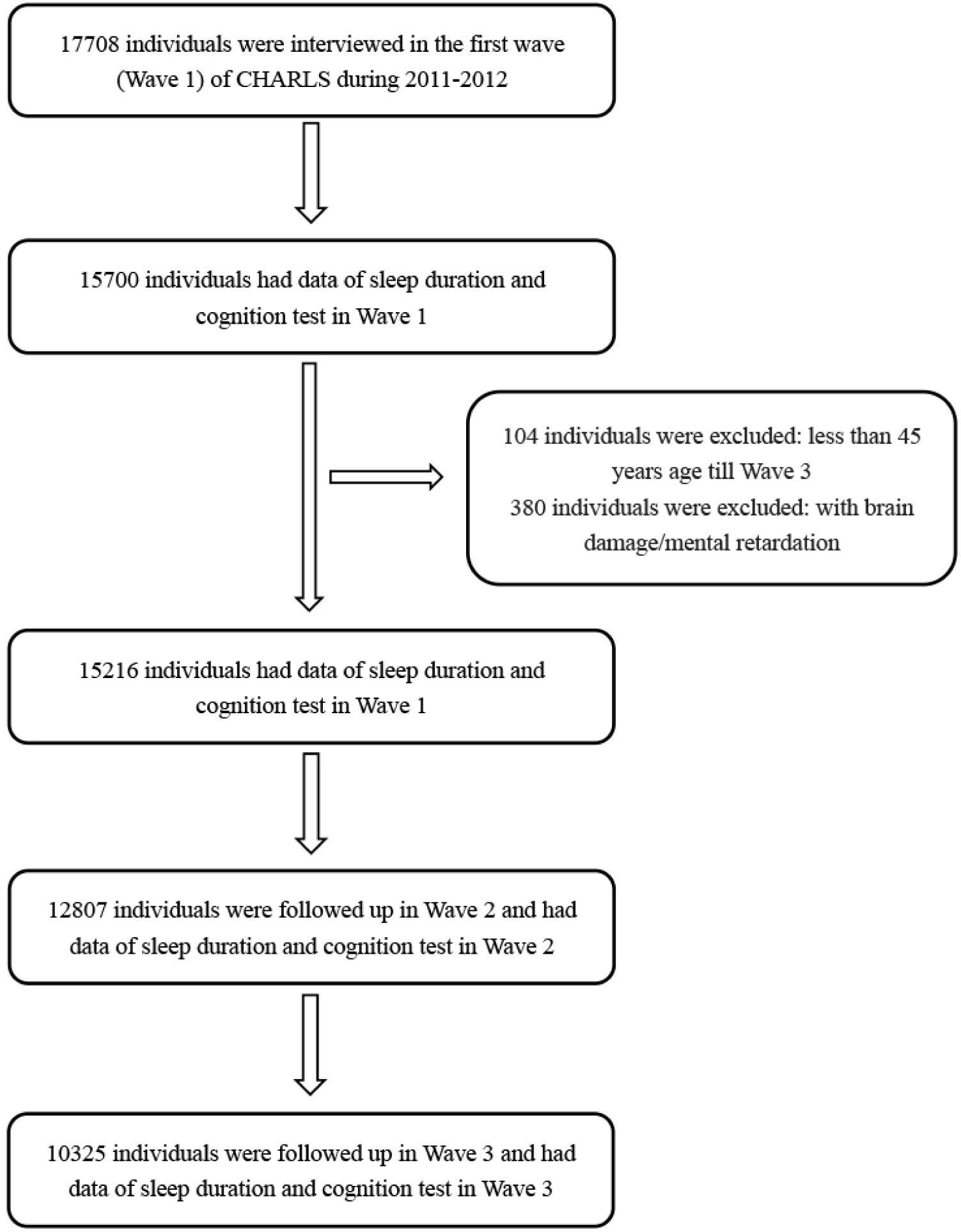
Flow chart of the sample selection and exclusion criteria.

### 2.2 Assessment of cognitive function

The cognitive dimensions of memory, visuospatial abilities, orientation, attention and calculation were assessed, through three tests: episodic memory, figure drawing, and Telephone Interview of Cognitive Status (TICS). Serving as the primary outcome, the global cognition score was the sum of the three test scores. The global cognition score could range from 0 to 21.

In the episodic memory test, the individuals were asked to recall the words immediately (immediate recall) and 5 minutes later (delayed recall) after interviewers read 10 Chinese nouns to them. The episodic memory score was the average score of the immediate recall and delayed recall tests and could range from 0 to 10^12^. This test assessed individuals’ memory.

In the figure-drawing test, the individuals were shown a picture and asked to redraw it. Those who succeeded in drawing got a score of 1. If failed, they got a score of 0. This test examined visuospatial abilities.

The TICS test was based on selected questions from the TICS battery, a well-established measure of one’s ability related to orientation, attention and calculation. In this test, the participants were asked to repeatedly subtract 7 from 100 and to identify the date, season, and day of the week. The TICS scores could range from 0 to 10^13^.

### 2.3 Assessment of sleep duration

Nocturnal sleep duration was obtained by the following questions, “During the past month, how many hours of actual sleep did you get at night (average hours for one night)”? The data was accurate to 0.5 h.

### 2.4 Potential Confounders

The potential confounders included age, gender, education, marital status, residential area, cigarette smoking, alcohol drinking, depression, instrumental activities of daily living (IADLs), use of tranquilizers, and comorbidities.

596 participants in our study missed data on body mass index (BMI). To maximize our sample size, we did not take BMI into account^14^. IADLs could range from 0 to 5 and reflect functional status^15^. Depression was classified as “yes” and “no”, using the 10-item Center for Epidemiologic Studies Short Depression Scale (CES-D-10). This score can range from 0 to 30, and the cut-off point for depression was 12^16^. The comorbidities included hypertension, dyslipidaemia, history of stroke, history of heart diseases^17^.

### 2.5 Statistical Analysis

Demographic characteristics were shown as the mean ± SD (standard deviation) or frequency (percentage). Associations between participants’ characteristics and baseline sleep duration were examined using ANOVA or the Pearson Chi-square test (Table 1).

**Table 1.** Demographic and health characteristics of the study population in Wave 1^*^

|  | <6 h (n=4453) | 6-8 h (n=9508) | >8 h (n=1255) | All (n=15216) | *P*<sup>a</sup> | *P*<sup>b</sup> |
| --- | --- | --- | --- | --- | --- | --- |
| Continuous variables |  |  |  |  |  |  |
| age | 61.0 (10.0) | 58.1(9,5) | 59.7(10.7) | 59.1 (9.8) | <0.001 | <0.001 |
| BMI | 23.0(3.6) | 23.6(3.6) | 23.2(3.6) | 23.4 (3.7) | <0.001 | <0.001 |
| Number of IADLs | 0.3(0.9) | 0.2(0.6) | 0.3(0.9) | 0.2 (0.7) | <0.001 | <0.001 |
| Categorical Variables, n (%) |  |  |  |  |  |  |
| Males | 1889(42.4) | 4711(49.6) | 587(46.9) | 7187 (47.3) | <0.001 | 0.073 |
| Education |  |  |  |  | <0.001 | <0.001 |
| Illiterate | 1492(33.5) | 2153(26.7) | 415(33.1) | 4060 (26.7) |  |  |
| Primary school | 1872(42.1) | 3667(38.6) | 513(40.9) | 6052 (39.8) |  |  |
| Middle school | 732(16.5) | 2211(23.3) | 222(17.7) | 3165 (20.8) |  |  |
| High school and above | 354(8.0) | 1470(15.5) | 105(8.4) | 1929 (12.7) |  |  |
| Marital status |  |  |  |  | <0.001 | <0.001 |
| Married | 3742(84.0) | 8566(90.1) | 1085(86.5) | 13393 (88.0) |  |  |
| Other status | 711(16.0) | 959(9.9) | 170(13.5) | 1823 (12.0) |  |  |
| Residential area |  |  |  |  | <0.001 | <0.001 |
| Urban | 847(19.1) | 2522(26.6) | 205(16.4) | 3574 (23.5) |  |  |
| Rural | 3600(81.0) | 6975(73.4) | 1047(83.6) | 11622 (76.5) |  |  |
| Depression | 301(6.8) | 654(6.9) | 118(9.5) | 1073 (7.1) | 0.795 | 0.001 |
| Use of tranquilizers | 25(0.6) | 20(0.2) | 4(0.0) | 49 (0.3) | <0.001 | 0.444 |
| Current smoker | 1660(37.2) | 3799(40.0) | 467(37.2) | 5926 (39.0) | 0.001 | 0.062 |
| Current drinker | 1040(23.4) | 2467(26.0) | 276(22.0) | 3783 (24.9) | 0.003 | 0.003 |
| Hypertension | 1187(26.7) | 2193(22.9) | 329(25.5) | 3688 (24.2) | <0.001 | 0.044 |
| Dyslipidaemia | 447(10.0) | 848(8.9) | 103(8.2) | 1397 (9.2) | 0.032 | 0.411 |
| History of heart disease | 680(15.3) | 1041(11.1) | 118(9.4) | 1839 (12.1) | <0.001 | 0.097 |
| History of stroke | 122(2.7) | 163(1.7) | 27(2.2) | 312 (2.1) | <0.001 | 0.269 |

\*Three groups were categorized by the sleep duration in Wave 1.
<sup>a</sup> *P* value for participants slept <6 h in Wave 1 compared with those slept 6-8 h.
<sup>b</sup> *P* value for participants slept >8 h in Wave 1 compared with those slept 6-8 h.

We used generalized additive models (GAM) to examine the inverted-U shaped association between baseline sleep duration and cognitive function in Wave 1 (Figure 2) and Wave 3 (Figure 3). GAM is “an extension of the generalized linear model which allows the evaluation for the curvilinear relationship of the outcome and the predictors”. The effective degree of freedom (EDF) output by GAM reflects the degree of curvature. An EDF = 1 means linear relation, while an EDF > 1 means more complex relations. Individuals whose sleep duration was out of mean ± 2SD were deleted temporarily^18^. Model 0 used global cognition score as a continuous response and a smoothing spline function of sleep duration as a univariable predictor. In model 1, the predicted variables were age, gender and smooth(sleep duration). The predicted smooth functions along with the 95% confidence intervals were plotted^19^.

**Figure 2.**
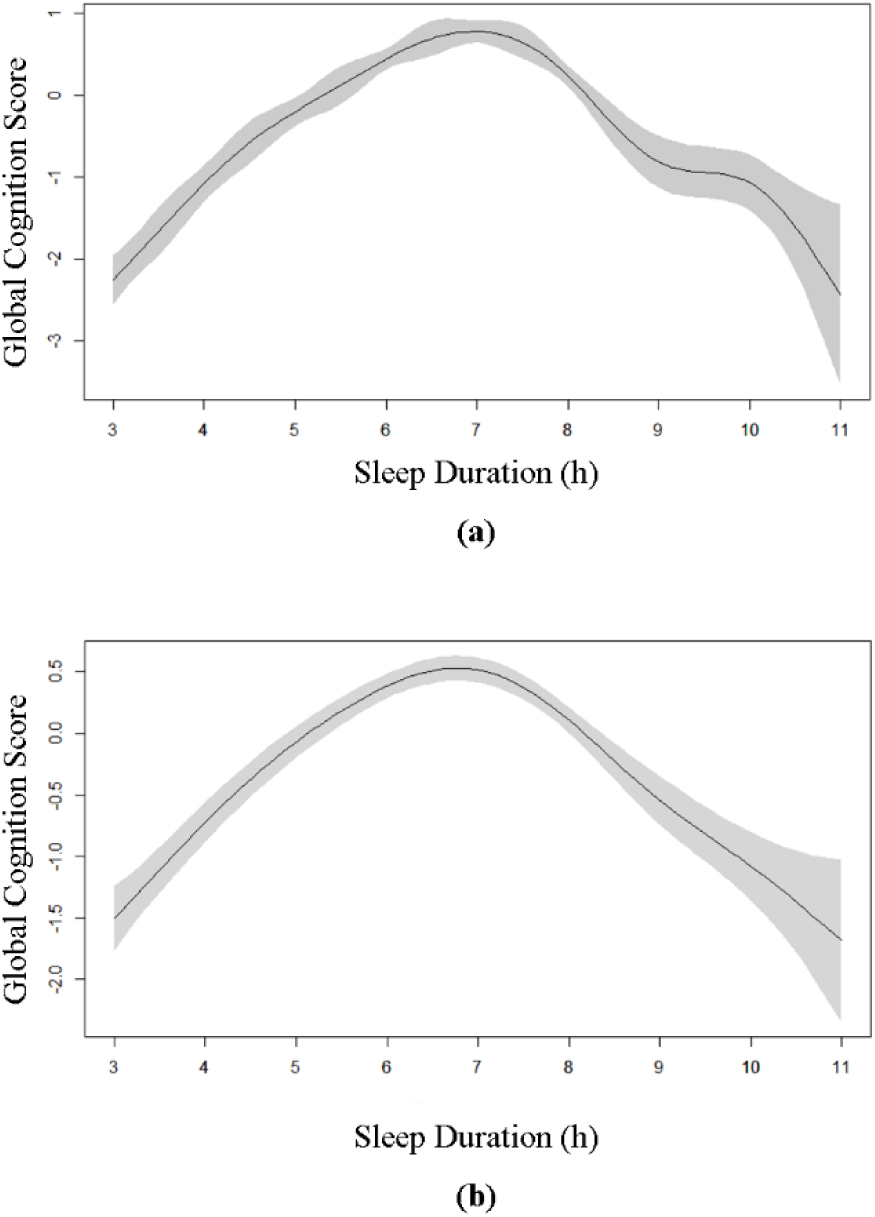
Plots of estimated smooth function of sleep duration in Wave 1 with 95% confidence intervals for the GAM when the response variable was global cognition in Wave 1. (A) Model 0 showed univariate smooth function of sleep duration (EDF = 7.44, *p* < 0.001). (B) Model 1 presented multivariable smooth function of sleep duration, adjusted for age and gender (EDF = 4,74, *p* < 0.001).

**Figure 3.**
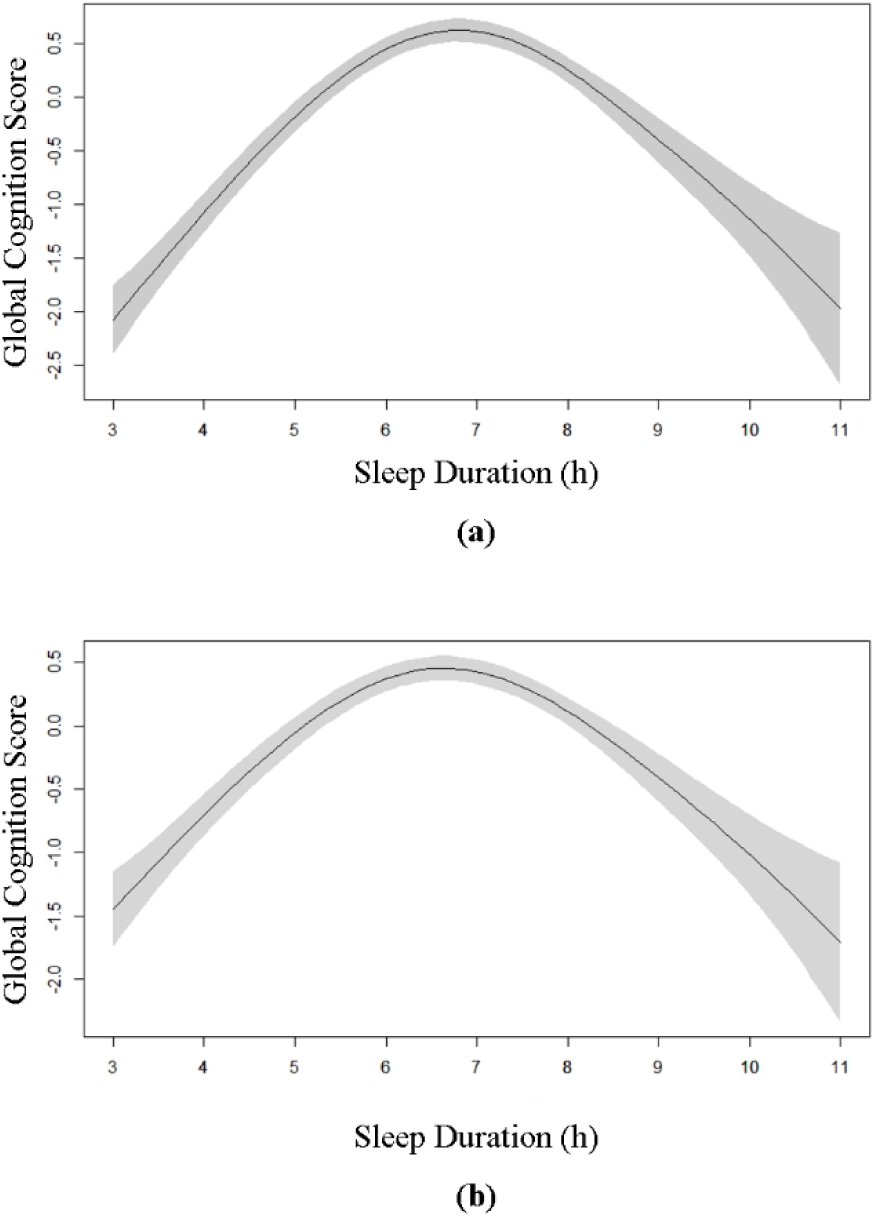
Plots of estimated smooth function of sleep duration in Wave 1 with 95% confidence intervals for the GAM when the response variable was global cognition in Wave 3. (A) Model 0 showed univariate smooth function of sleep duration (EDF = 3.71, *p* < 0.001). (B) Model 1 presented multivariable smooth function of sleep duration, adjusted for age and gender (EDF = 4.58, *p* < 0.001).

We categorized sleep duration into three patterns: short sleep duration (SSD), <6 h; moderate sleep duration (MSD), 6-8h; long sleep duration (LSD), >8h. All previous literature chose 6 h as the lower cut-off point. For the mean sleep duration of our participants was 6.4 h and our participants’ optimal sleep duration for global cognition was about 7 h, we chose 8 h as the upper cut-off point.

Generalized estimation equations (GEE) were used to extend the GAM for further analysis of the longitudinal association between baseline sleep duration and cognitive function over a period of 4 years. GEE accounted for between-participant variation and within-participant correlation of repeated outcomes. Time was defined as a continuous variable, measured as years from baseline. In model 1 and model 2, we calculated the estimates of sleep duration (Table 2), to examine the association between baseline sleep duration and cognition. In model 3, we calculated the estimates of interaction of time and sleep duration, to study the rate of cognitive decline^20,21^.

**Table 2.** Associations between baseline sleep duration and longitudinal global cognition by generalized estimating equations (GEE)

| | Model 1 $\beta$(SE) | Model 2 $\beta$(SE) | Model 3 $\beta$(SE) |
| --- | --- | --- | --- |
| **Categorical trend** |  |  |  |
| SSD (<6 h) | -0.94 (0.06)*** | -0.53 (0.06)*** |  |
| MSD (6-8 h) | Ref. | Ref. |  |
| LSD (>8 h) | -1.05 (0.11)*** | -0.59 (0.09)*** |  |
| SSD*time |  |  | 0.02 (0.02) |
| MSD*time |  |  | Ref. |
| LSD*time |  |  | -0.03 (0.04) |
| **Linear trend for sleep duration $\leq 7$ h** |  |  |  |
| Duration | 0.48 (0.03)*** | 0.28 (0.02)*** |  |
| Duration*time |  |  | 0.004 (0.009) |
| **Linear trend for sleep duration $\geq 7$ h** |  |  |  |
| Duration | -0.47 (0.02)*** | -0.28 (0.04)*** |  |
| Duration*time |  |  | -0.007 (0.015) |

Abbreviations: SSD, short sleep duration; MSD, moderate sleep duration; LSD, long sleep duration.
\*\*\* $P<0.001$ , $\beta$ : unstandardized beta coefficient, SE: standard error.

To analyse associations between change in sleep duration and cognition, we used generalized linear models (GLM), after adjustment for potential confounders.

As discussed above, the global cognition score was the primary outcome. Associations with the three cognition tests included in the global cognition score were examined in complementary analyses and shown in supplementary material. GAM was carried out using R-3.4.3. Other statistical analyses were performed by SAS version 9.4 (SAS Institute Inc., Cary, North Carolina, USA).

## 3 Results

### 3.1 Demographic and health characteristics of the study population in Wave 1

In wave 1, the mean age of all participants was 59.1 ±9.8 years; 47.3% of the participants were male, 76.5% of them were from rural areas, and 88.0% of them were married. With regard to education, 73.3% of the participants attended primary school and above. The mean ±SD nocturnal sleep duration was 6.4 ±1.9 hours per night. At baseline, 62.5% of the participants were moderate sleepers (6-8 h), 29.2% of them were short sleeps (<6 h), while only 8.2% of them were long sleepers (>8 h). 0.3% individuals used tranquilizers.

In wave 1, the mean age of moderate sleepers was 2.9 years younger than short sleepers and 1.6 years younger than long sleepers (Table 1). Moderate sleepers were more likely to be male, live in urban areas, have better education, and get married. Moderate sleepers had a slightly higher BMI and lower number of IADLs, and they more often smoke and drunk. Long sleepers were more likely to suffer from depression. Short sleepers were more likely to have a history of hypertension, dyslipidemia, heart disease and stroke, while moderate sleepers tended to be healthier (Table 1).

### 3.2 Associations between baseline sleep duration and cognitive function

#### 3.2.1 Baseline sleep duration and cognitive function in Wave 1 and Wave3

The result of GAM for global cognition score and sleep duration indicated a non-linear fit in Wave 1(EDF = 7.44, *P*<0.001, Figure 2) and Wave 3(EDF = 3.71, *P* < 0.001, Figure 3). We found an inverted-U shaped cross-sectional association of sleep duration and global cognition score. The highest global cognition scores in Wave 1 and in Wave 3 were both probably observed among those sleeping 6-8 h per night in Wave 1. Remarkably, people sleeping 3 h or 11 h showed similar mean scores whichever wave.

#### 3.2.2 Baseline sleep duration and longitudinal cognitive function in three Waves

Short sleepers in Wave 1 had lower global cognition scores (β=-0.94, *P*<0.001 for model 1 and β=-0.53, *P*<0.001 for model 2), compared with moderate sleepers (Table 2). The specific domains were all three functions: episodic memory, figure drawing, and TICS (supplementary material). Expressed in age equivalents and compared to moderate sleep duration group, the effect of short sleep was equivalent to being 9 (model 1) or 5 (model 2) years older for global cognition score (data not shown). Longer sleepers in Wave 1 had lower global cognition scores (β=-1.05, *P*<0.001 for model 1 and β=-0.59, *P*<0.001 for model 2), compared to participants with a moderate sleep duration. The specific domains were all three tests. Expressed in age equivalents and compared to moderate sleep duration group, the effect of long sleep time was equivalent to being 11 (model 1) or 6 (model 2) years older for global cognition score.

Linear trends were also significant. For participants slept ≤7 h in Wave 1, lower sleep duration was associated with lower scores in global cognition (β=0.48, *P*<0.001 for model 1 and β=0.28, *P*<0.001 for model 2) and all three tests. For participants slept ≥7 h in Wave 1, higher sleep duration was associated with lower scores in global cognition (β=-0.47, *P*<0.001 for model 1 and β=-0.28, *P*<0.001 for model 2) and all three tests.

The sleep-duration-by-time reaction was not significant (*p* > 0.10 for model 3), showing no significant association between sleep duration in Wave 1 and decline rate of global cognition score over a period of 4 years (Table 2).

Model 1: adjusted for age and gender.

Model 2: adjusted for Model 1+ education, marital status, residential area, depression, IADLs, use of tranquilizers, smoking, drinking, hypertension, dyslipidaemia, heart disease and stroke.

Model 3: adjusted for age and gender.

### 3.3 Associations between change in sleep duration and cognitive function

#### 3.3.1 Participants slept 6-8 h in Wave 1

In the analysis of the effect of changes in sleep duration from Wave 1 to Wave 2 (Table 3) and from Wave 1 to Wave3 (Table 4) among moderate sleepers (6-8 h) in Wave 1, we also observed an inverted U-shaped association. Changes in sleep duration that increased (β=-1.30, *P*<0.001 for model 1, β=-0.83, *P*<0.001 for model 2 and β=-0.63, *P*<0.001 for model 3) or decreased (β=-0.87, *P*<0.001 for model 1, β=-0.52, *P*<0.001 for model 2 and β=-0.37, *P*<0.001 for model 3) ≥2 h over time were associated was lower global cognition scores than no change group (Table 3). The effect of ≥2 h change in Wave 2 was approximately equivalent to 9 years of cognitive aging in model 1, model 2, and model 3. These associations were consistent across all three cognition tests for model 1, model 2 and model 3 (supplementary material).

**Table 3.** Associations between change in sleep duration and global cognition in Wave 2 among participants slept 6-8 h at baseline^a^

| | Decreased by $\geq 2$ h | Decreased by 0.5-1.5 h | No change | Increased by 0.5-1.5 h | Increased by $\geq 2$ h |
| --- | --- | --- | --- | --- | --- |
| Number of subjects | 1487 | 1587 | 1690 | 1092 | 555 |
| Mean (SD) score | 7.79 (3.33) | 8.76 (3.18) | 8.80 (3.25) | 8.43 (3.28) | 7.25 (3.55) |
| Model 1 | -0.87 (0.11)***,b | -0.03 (0.11) | Ref. | -0.32 (0.12)** | -1.30 (0.16)*** |
| Model 2 | -0.52 (0.10)*** | -0.02 (0.10) | Ref. | -0.14 (0.11) | -0.83 (0.14)*** |
| Model3 | -0.37 (0.09)*** | -0.01 (0.09) | Ref.. | -0.09 (0.10) | -0.63 (0.13)*** |

<sup>a</sup> Using generalized linear models (GLM).
<sup>b</sup> Unstandardized beta coefficient (standard error), for all such values.
\*\* $P < 0.01$ , \*\*\* $P < 0.001$ .

**Table 4.**
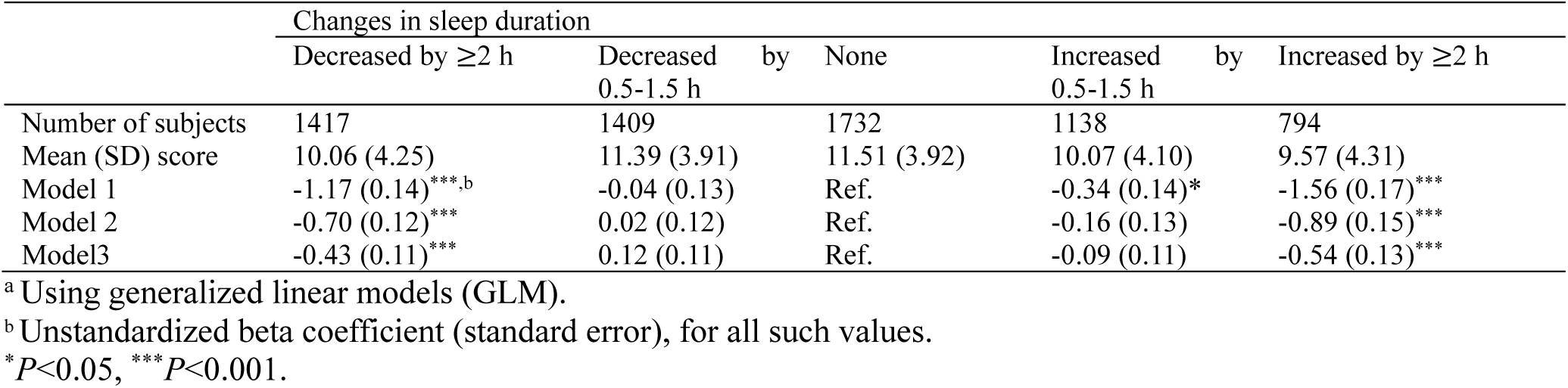
Associations between change in sleep duration and global cognition in Wave 3 among participants slept 6-8 h at baseline ^a^

While analyzing the effect of change from Wave 1 to Wave 3, we found the association between changes in sleep duration and global cognition remained unchanged (Table 4). This association was across all three tests for model 1. In model 2 and model 3, figure drawing test had no significance, while episodic memory test and TICS test remained significant.

We also studied short sleepers in Wave 1, reaching a similar result that ≥2 h change in Wave 2 or Wave 3 was associated with lower global cognition scores. For long sleepers in Wave 1, the effect made no difference (data not shown).

Model 1: adjusted for age and gender.

Model 2: adjusted for Model 1+ education, marital status, residential area, depression, IADLs, use of tranquilizers, smoking, drinking, hypertension, dyslipidaemia, heart disease and stroke.

Model 3: adjusted for Model 2 + baseline global cognition score

#### 3.3.2 Participants slept <6 h in Wave 1

The effect of sleep change on participants who slept <6 h in Wave 1 was studied by sub-group analysis (Table 5). Compared to “No change” group, “Excessive” group had lower global cognition in Wave 3 (β=-1.91, *P*<0.001 for model 1, β=-0.94, *P*<0.001 for model 2 and β=-0.53, *P*<0.05 for model 3). The specific domains included all three tests. “Benefit 1” group showed no significant difference. “Benefit 2” group had higher global cognition score (β=0.55, *P*<0.01 for model 1, β=0.54, *P*<0.01 for model 2 and β=0.38, *P*<0.05 for model 3). The effect of “Benefit 2” was approximately equivalent to 4-10 years aging. The specific domains were figure drawing and TICS, while episodic memory had no significance (supplementary material).

**Table 5.** Associations between change in sleep duration and global cognition in Wave 3 among participants slept <6 h or >8 h at baseline^a^

|  | Type of change in sleep duration<sup>c</sup> |  |  |  |
| --- | --- | --- | --- | --- |
|  | Excessive | No change | Benefit 1 | Benefit 2 |
| **SSD (<6 h) in Wave 1** |  |  |  |  |
| Number of subjects | 224 | 1231 | 937 | 602 |
| Mean (SD) score | 7.23 (4.42) | 9.39 (4.36) | 9.27 (4.23) | 10.35 (4.03) |
| Model 1 | -1.91 (0.30)<sup>***,b</sup> | Ref. | -0.31 (0.18) | 0.55 (0.20)<sup>**</sup> |
| Model 2 | -0.94 (0.26)<sup>***</sup> | Ref. | -0.22 (0.15) | 0.54 (0.17)<sup>**</sup> |
| Model 3 | -0.53 (0.23)<sup>*</sup> | Ref. | -0.21 (0.14) | 0.38 (0.15)<sup>*</sup> |
| **LSD (>8 h) in Wave 1** |  |  |  |  |
| Number of subjects | 232 | 64 | 217 | 328 |
| Mean (SD) score | 8.19 (4.58) | 8.77 (4.22) | 9.47 (4.32) | 10.47 (4.24) |
| Model 1 | -0.68 (0.58) | Ref. | 0.22 (0.54) | 0.80 (0.54) |
| Model 2 | -1.38 (0.52)<sup>**</sup> | Ref. | -0.72 (0.50) | -0.34 (0.52) |
| Model 3 | -1.17 (0.45)<sup>**</sup> | Ref. | -0.72 (0.46) | -0.43 (0.46) |

Abbreviations: SSD, short sleep duration; MSD, moderate sleep duration; LSD, long sleep duration.
<sup>c</sup> Type of change was decided by patterns of sleep duration in Wave 2 and Wave3
For SSD in Wave 1: Excessive, as long as one LSD in each wave; No change, remained SSD in two waves; Benefit 1, remained SSD in one wave and MSD in another; Benefit 2, MSD in two waves.
For LSD in Wave 1: Excessive, as long as one SSD in each wave; No change, remained LSD in two waves; Benefit 1, remained LSD in one wave and MSD in another; Benefit 2, MSD in two waves.
<sup>a</sup> Using generalized linear models (GLM).
<sup>b</sup> Unstandardized beta coefficient (standard error), for all such values.
<sup>\*</sup> $P<0.05$ , <sup>\*\*</sup> $P<0.01$ , <sup>\*\*\*</sup> $P<0.001$ .

#### 3.3.3 Participants slept >8 h in Wave 1

The effect of sleep change on participants who slept >8 h in Wave 1 was also shown in Table 5. Compared to “No change” group, “Excessive” group had lower global cognition in Wave 3 (β=-1.38, *P*<0.01 for model 2 and β=-1.17, *P*<0.01 for model 3). The specific domain was episodic memory. Participants in “Benefit 1” group and “Benefit 2” group had higher global cognition scores. However, there was no significant difference after adjusted for confounders.

Model 1: adjusted for age and gender.

Model 2: adjusted for Model 1+ education, marital status, residential area, depression, IADLs, use of tranquilizers, smoking, drinking, hypertension, dyslipidaemia, heart disease and stroke.

Model 3: adjusted for Model 2 + baseline global cognition score

## 5 Discussion

### 5.1 Synopsis of findings

So far, this was the largest and latest study examining the longitudinal association between self-reported sleep duration and cognitive function^3^.Using GAM and GEE models, our study indicated an inverted-U shaped association between baseline sleep duration and global cognition over a period of 4 years, among Chinese participants. The affected domains included all tests: episodic memory, figure drawing and TICS. However, baseline sleep duration would not affect the rates of cognitive change. For moderate sleepers, ≥2 h change in sleep duration was significantly associated with lower global cognition and all three tests. Relative to those whose sleep duration remained unchanged, a move from SSD to LSD was associated with lower global cognition, included all three tests, and a move from LSD to SSD was associated with lower episodic memory scores. For short sleepers, a consistent change to MSD was associated with high global cognition scores, equivalent to 4-10 years of cognitive aging. The improved domains were figure drawing and TICS.

A change from LSD to MSD had no significant effect. However, only 8.2% of the participants slept LSD at baseline and 0.6% of the participants remained LSD from Wave 1 to Wave 3.

### 5.2 Comparison with other studies

The most important finding of our study was that we challenged the previous ideas that increased or decreased sleep duration would lead to lower cognition. Up to data, a total of 9 studies have examined the effect of change in sleep duration on cognitive function or risk of dementia. 5 studies linked increased sleep duration to lower cognition or higher risk of dementia^8,10,22-24^, 1 study linked decreased sleep duration to higher risk of AD^25^, and 1 study found no association^26^. While 2 studies reported both increase and decrease in sleep duration was associated with lower cognition^9,27^.

Using sub-group analysis, we discovered that a change to MSD was associated with better cognition, while a deviation from MSD or excessive change would lead to worse cognition. The following reasons could explain why conclusions of previous studies differed from ours: First, for moderate sleepers, our finding was in accordance with those studies’ that a specific degree of change was harm to cognition. Second, for short sleepers, the positive effect of change to MSD was covered by the negative effect of excessive change to LSD. Third, for long sleepers, the negative effect of the ≥2 hours decrease in sleep duration was caused by the change from LSD to SSD, while change from LSD to MSD was not associated with worse cognition.

Table 6 showed studies focused on change in sleep duration and used sub-group analysis. Based on the sleep duration at baseline, participants were divided into 3 groups: SSD, MSD, and LSD. The participants in the Chinese Longitudinal Healthy Longevity Survey had a mean age of 80 y, and 30% of them died during a 3-year follow-up. The age might restrict them from finding the association. Furthermore, the sleep pattern of their participants differed widely from other studies, that more (11.5% vs. <1%) participants remained LSD^24^. The sleep duration in the Ohsaki Cohort 2006 study was recorded in 1994 and 2006. The human’s sleep duration declines as age grows older^28^. The decline in sleep duration caused by the long follow-up time would affect the result^23^. The Whitehall Study had the relatively best study design, nonetheless, they did not find significance among people with SSD or have enough samples of LSD^9^. Remarkably, the findings in the 3 studies listed in Table 6 were not contradictory to our findings. These studies just did not reach our findings and repeated the previous conclusion that increased or decreased sleep duration would cause lower cognition. Last but not least, in our study, a change from LSD to MSD had no effect. The Chinese Longitudinal Healthy Longevity Survey reported a change from LSD to MSD could decreased the risk of dementia, and it could be a supplementary to our result.

**Table 6.**
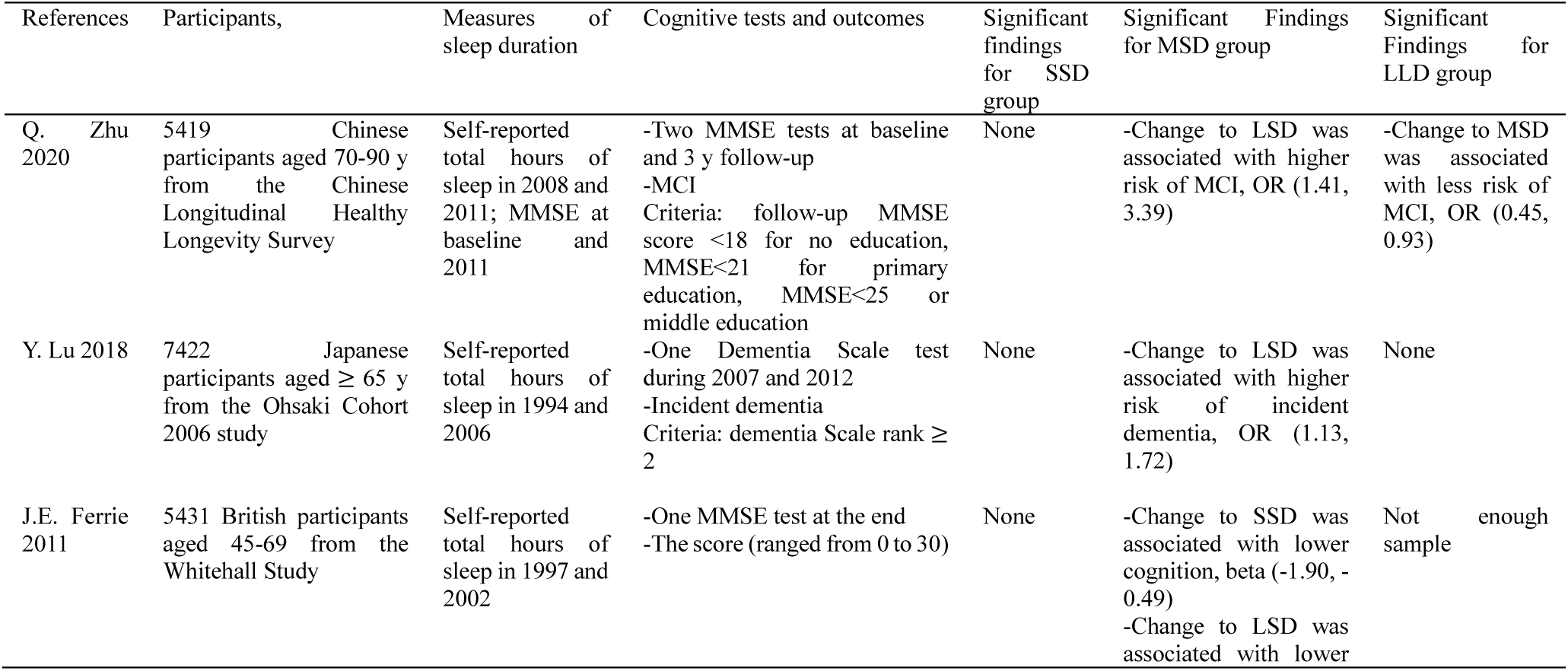
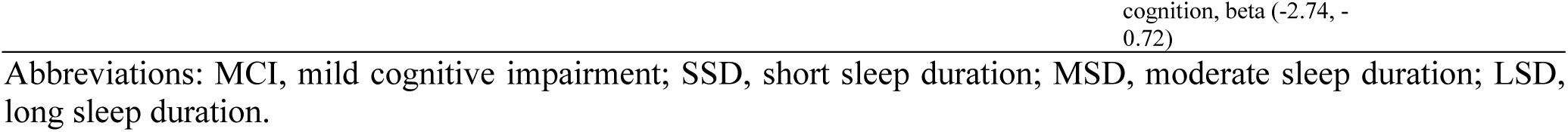
Summary of studies that linked changes in sleep duration with cognitive function or dementia, using sub-group analysis

Whether sleep duration affected the rate of cognitive change was controversial^14,21,27,29-32^. Some reported long sleep duration at baseline would lead to faster decline of cognition, while some reported that there was no association. Using GEE, we did not find association between baseline sleep duration and cognitive decline.

### 5.5 Possible mechanisms explaining improvement in sleep duration and cognitive function

The circadian rhythm was composed of clock genes in almost every cell of human body and part of the neuro-endocrine system. It provides humans with the adaption to the earth rotation every 24 hours. There was a bidirectional relationship between sleep duration, circadian rhythm and cognitive function^6^. Short or long sleep duration could disrupt circadian rhythm^5^. Numerous studies pointed to short or long sleep duration and disruption of circadian rhythm as a risk factor for neurodegenerative diseases, including dementia.^4,33^ Few clinical studies reported that restoration of circadian rhythm might reduce the deterioration of human’s cognitive function, by using controlled light exposure or injection of melatonin, an endocrine hormone associated with circadian rhythm^34-37^. In an animal study, scientists improved the sleep duration of mice carrying the Huntington’s Disease (HD) mutation, slowing down the cognitive decline of mice and reversed dysregulation of their circadian rhythm.^38,39^ These studies suggested that a change from LSD to MSD might restore the circadian rhythm and thus led to better cognitive function in our participants. Inversely, excessive change and deviation from MSD correlated with disruption of circadian rhythm, which was associated with worse cognitive function.

Furthermore, other mechanisms were discovered for explanation of the cross-sectional association between sleep duration and cognitive function, such as inflammatory markers, sleep apnea and sleep fragmentation. Nonetheless, whether change in sleep duration would affect these mediators was not clear.

### 5.4 Advantages and limitations

A strength of our study was its large and national-representative sample of middle-aged and older Chinese, who were enrolled in a prospective and up-to-date study. And, the robustness of the CHARLS survey allowed us to adjust for multiple confounders.

Our study also had several limitations. First, measurement of sleep duration was self-reported, which would be influenced by recall bias. *C*.*L. Jackson 2018* studied the differences between self-reported and objectively measured sleep duration among several ethics. Compared with whites (73 min, 95% CI: 67-79), blacks and Hispanics, Chinese had the lowest bias (49 min, 95% CI: 37-61)^40^. Plenty of cohorts found association between self-reported sleep duration and unhealthy outcomes, making our research convincible^41^. Most importantly, self-reported measurement is cheap and practicable, increasing the applicable of our results for health education and general research. Second, sleep-disordered breathing (SDB), which was no designed in ours study, was viewed to affect cognition. ^6,42,43^. A recent study assessed 5247 participants with in-home polysomnography and found no association between cognitive function and SDB assessments, including Epworth Sleepiness Scale and Respiratory Event Index^7^. Third, 15.9% of participants in Wave 1 lost to follow up in Wave 2 and 19.4% lost to follow up in Wave 3. This might affect our result.

### 5.5 For health education

For short sleepers, a consistent change to moderate sleep duration correlated with better cognition. For long sleepers, there was no need to reduce sleep duration. Excessive changes over the moderate sleep duration or deviation from the moderate was associated with lower cognition.

## Data Availability

The data used in this manuscript from the China Health and retirement Longitudinal Study (CHARLS). We applied the permission for the data access (http://charls.pku.edu.cn/zh-CN) and got the access to use it. Prof. Yaohui Zhao (National School of Development of Peking University), John Strauss (University of Southern California), and Gonghuan Yang (Chinese Center for Disease Control and Prevention) are the principle investigators.

## 7 Acknowledgments

We appreciated the China Center for Economic Research, the National School of Development of Peking University for providing the data.

## 8 Author Contributions

Jianian Hua contributed conception and design of the study. Yueping Shen organized the database. Jianian Hua performed the statistical analysis. Jianian Hua wrote the first draft of the manuscript. Yueping Shen and Hongpeng Sun reviewed the manuscript. All authors approved the final version of the paper.

## 9 Ethics Statement

Each participant included in this study signed a written informed consent form before taking the survey. Ethics approval for the data collection in the CHARLS was obtained from the Biomedical Ethics Review Committee of Peking University (IRB00001052-11015). We confirm that all methods were performed in accordance with the relevant guidelines and regulations.

## 10 Disclosure Statement

Financial Disclosure: none. Non-financial Disclosure: none.

## 12 Funding

This work was supported by the National Natural Science Foundation of China (project number 81973143) and the Priority Academic Program Development of Jiangsu Higher Education Institutions (PAPD).

## Abbreviation

SSD: short sleep duration
MSD: moderate sleep duration
LSD: long sleep duration.

## Notes

### Competing Interest Statement

The authors have declared no competing interest.

